# Multi-site Technical Performance and Concordance of Optical Genome Mapping: Constitutional Postnatal Study for SV, CNV, and Repeat Array Analysis

**DOI:** 10.1101/2021.12.27.21268432

**Authors:** M. Anwar Iqbal, Ulrich Broeckel, Brynn Levy, Steven Skinner, Nikhil Sahajpal, Vanessa Rodriguez, Aaron Stence, Kamel Awayda, Gunter Scharer, Cindy Skinner, Roger Stevenson, Aaron Bossler, Peter L. Nagy, Ravindra Kolhe

## Abstract

**Background:** The standard of care (SOC) cytogenetic testing methods, such as chromosomal microarray (CMA) and Fragile-X syndrome (FXS) testing, have been employed for the detection of copy number variations (CNVs), and tandem repeat expansions/contractions that contribute towards a sizable portion of genetic abnormalities in constitutional disorders. However, CMA is unable to detect balanced structural variations (SVs) or determine the precise location or orientation of copy number gains. Karyotyping, albeit with lower resolution, has been used for the detection of balanced SVs. Other molecular methods such as PCR and Southern blotting, either simultaneously or in a tiered fashion have been used for FXS testing, adding time, cost, and complexity to reach an accurate diagnosis in affected individuals. Optical genome mapping (OGM), innovative technology in the cytogenomics arena enables a direct, high-resolution view of ultra-long DNA molecules (more than 150 kbp), which are then assembled *de novo* to detect germline SVs ranging from 500 bp insertions and deletions to complex chromosomal rearrangements. The purpose of this study was to evaluate the performance of OGM in comparison to the current SOC methods and assess the intra- and inter-site reproducibility of the OGM technique. We report the largest retrospective dataset to date on OGM performed at five laboratories (multi-site) to assess the robustness, QC performance, and analytical validation (multi-operator, and multi-instrument) in detecting SVs and CNVs associated with constitutional disorders compared to SOC technologies.

**Methods:** This multi-center IRB-approved, double-blinded, study includes a total of 331 independent flow cells run (including replicates), representing 202 unique retrospective samples, including but not limited to pediatric-onset neurodevelopmental disorders. This study included affected individuals with either a known genetic abnormality or no known genetic diagnosis. Control samples (n=42) were also included. Briefly, OGM was performed on either peripheral blood samples or cell lines using the Saphyr system. The OGM assay results were compared to the human reference genome (GRCh38) to detect different types of SVs (CNV, insertions, inversions, translocations). A unique coverage-based CNV calling algorithm was also used to complement the SV calls. Analysis of heterozygous SVs was performed to assess the absence of heterozygosity (AOH) regions in the genome. For specific clinical indications of FSHD1 and FXS, the EnFocus FXS and FSHD1 tools were used to generate the region-specific reports. OGM data was analyzed and visualized using Access software (version 1.7), where the SVs were filtered using an OGM specific internal control database. The samples were analyzed by laboratory analysts at each site in a blinded fashion using ACMG guidelines for SV interpretation and further reviewed by expert geneticists to assess concordance with SOC testing results.

**Results:** Of the first 331 samples run between five sites, 99.1% of sample runs were completed successfully. Of the 331 datasets, 219 were assessed for concordance by the time of this publication; these were samples that harbored known variants, of which 214/219 were detected by OGM resulting in a concordance of 97.7% compared to SOC testing. 47 samples were also run in intra- and inter-site replicate and showed 100% concordance for pathogenic CNVs and SVs and 100% concordance for pathogenic *FMR1* repeat expansions.

**Conclusion:** The results from this study demonstrate the potential of OGM as an alternative to existing SOC methods in detecting SVs of clinical significance in constitutional postnatal genetic disorders. The outstanding technical performance of OGM across multiple sites demonstrates the robustness and reproducibility of the OGM technique as a rapid cytogenomics testing tool. Notably, OGM detected all classes of SVs in a single assay, which allows for a faster result in cases with diverse and heterogeneous clinical presentations. OGM demonstrated 100% concordance for pathogenic variants previously identified including *FMR1* repeat expansions (full mutation range), pathogenic D4Z4 repeat contractions (FSHD1 cases), aneuploidies, interstitial deletions, interstitial duplications, intragenic deletions, balanced translocations, and inversions. Based on our large dataset and high technical performance we recommend OGM as an alternative to the existing SOC tests for the rapid detection and diagnosis of postnatal constitutional disorders.

## Introduction

Constitutional disorders, including birth defects, pediatric and adult neurodevelopmental disorders warrant genetic/genomic testing for accurate disease diagnosis and clinical management. Specifically, neurodevelopmental disorders (NDDs), a group of disorders characterized by developmental deficits in cognition, behavior, language, and/or motor skills affects up to 17% of the pediatric population in the United States [**1**]. Given that NDDs are clinically and etiologically heterogeneous disorders [**2**], genetic testing has been employed globally since the 1970s. Different classes of SVs in the genome, such as aneuploidies, translocations, inversions, interstitial deletions/duplications, repeat expansions/contractions, and complex rearrangements contribute to a significant number of constitutional disorders [**3**]. Prior to 2010, G-banded karyotype was the recommended genetic testing method for individuals with unexplained neurodevelopmental disabilities, and other defects to detect balanced translocations, inversions, and other chromosomal rearrangements. However, karyotyping demonstrated a diagnostic yield of approximately 5% for these disorders [**4**]. In the mid-2000s, multiple studies showed the potential of chromosomal microarray analysis (CMA) as an effective clinical tool for the detection of whole-genome copy number variations (CNV). CMA can detect large CNVs and unbalanced rearrangements like karyotype analysis, and can additionally detect smaller duplications, deletions, and absence of heterozygosity, providing a diagnostic yield up to ∼20% [**5**], partially overlapping those found by karyotype analysis. Subsequently, CMA was recommended as the first-tier clinical diagnostic test by several medical organizations and societies (including the American College of Medical Genetics and Genomics [**6**], the American Academy of Neurology [**7**] and the American Academy of Pediatrics [**8, 9**] for individuals with developmental delay, intellectual disability, autism spectrum disorder (ASD), dysmorphic features, and multiple congenital anomalies. However, CMA has several technical limitations that include the inability to detect balanced structural rearrangements, decipher the orientation of duplicated segments of the genome, the limit of resolution restricted to few kilobases, and the inability to detect low-level mosaicism [**10**]. In addition to CMA as a front-line test, FXS testing is also recommended in the diagnostic workup for individuals with developmental delay/intellectual disability and ASD [**11**].

Recent advances in sequencing technology have enabled laboratories to evaluate and offer targeted panels, whole exome (ES), and genome sequencing (GS) for genetic testing of individuals with a clinical suspicion of NDDs. In a recent meta-analysis, ES has been shown to have a diagnostic yield of ∼35% and is currently offered by clinical laboratories as a reflex test after a non-diagnostic CMA [**12**]. Several groups have made attempts to detect SVs with ES and GS, but the repetitive sequences in the genome that facilitate the enrichment of SVs remain difficult to capture as a result of an innate limitation of the short read lengths and their failure to unambiguously map correctly. Moreover, CMA and ES cannot detect repeat expansion disorders; for example, the trinucleotide expansion of repeats of CGG within the *FMR1* gene causing FXS or D4Z4 repeat array contraction for the detection of facioscapulohumeral muscular dystrophy (FSHD1). Therefore, ancillary methods such as Southern blotting or PCR testing are required for repeat array analysis, and although these technologies have been in clinical use, they are time-consuming and have low accuracy (Southern blotting) or have a low dynamic range (PCR, NGS). Nevertheless, a cumulative diagnostic yield with all these technologies is estimated to be over 50% for individuals with developmental delay, intellectual disability and over 25% for individuals with primarily ASD [**10**]. Clearly, there is a need for better techniques that can detect the above-mentioned variant classes using fewer assays and also provide answers in a large number of cases that remain “negative” by current SOC technologies.

Optical genome mapping is one such technique, which is emerging as a next-generation cytogenomic tool as it enables a comprehensive cytogenomic analysis of the genome [**13**]. OGM, in its current iteration, is developed on the Saphyr system, and marketed by Bionano Genomics (San Diego, CA). OGM employs the imaging of ultra-long DNA molecules (>150 kbp) that are labeled at a unique 6 base-pair sequence motif (CTTAAG) that occurs throughout the entire genome. The images of the labeled DNA molecules are used to generate a *de novo* assembly that can be compared to a reference genome to identify all classes of SVs, such as deletions, duplications, balanced/unbalanced genomic rearrangements (insertions, inversions, and translocations), and repeat array expansions/contractions [**14**]. In addition, a separate coverage-based algorithm enables the detection of genome-wide copy number analysis (similar to a CMA), and the absence of heterozygosity (AOH) analysis. In the same assay, a concurrent or stepwise data analysis pipeline allows for sizing pathogenic CGG repeat expansions (FXS) as well as D4Z4 repeat contractions (FSHD1)[**15**]. Recently, in several evaluation/validation studies, OGM has demonstrated near-perfect concordance to standard-of-care testing [**16-17**]. Additionally, OGM has been validated and is clinically offered for FSHD1 testing [**18**]. Importantly, the OGM workflow is often more efficient than SOC test protocols, providing results within three-five days compared to average first-tier test turn-around times especially considering that many of those are non-diagnostic and require additional tiers of testing [**19-21**].

The aim of this double-blinded, multi-site, observational, IRB approved study was to evaluate and validate the performance of OGM compared to SOC technologies (CMA, karyotyping, Southern blotting, and PCR), in a large sample set of 331 sample runs containing 202 unique samples (made up of aneuploidies, intragenic and contiguous deletions, duplications, balanced and unbalanced translocations, inversions, isochromosomes, ring chromosomes, repeat expansions, repeat contractions, and more). This multi-site study assesses the performance of the OGM workflow by multiple operators, using multiple instruments (figure 1) to demonstrate the robustness and sensitivity of the technology. Consensus workflow and interpretation protocols were developed and validated to assess if OGM can replace the current standard-of-care technologies for the genetic testing of constitutional disorders.

**Figure 1.**
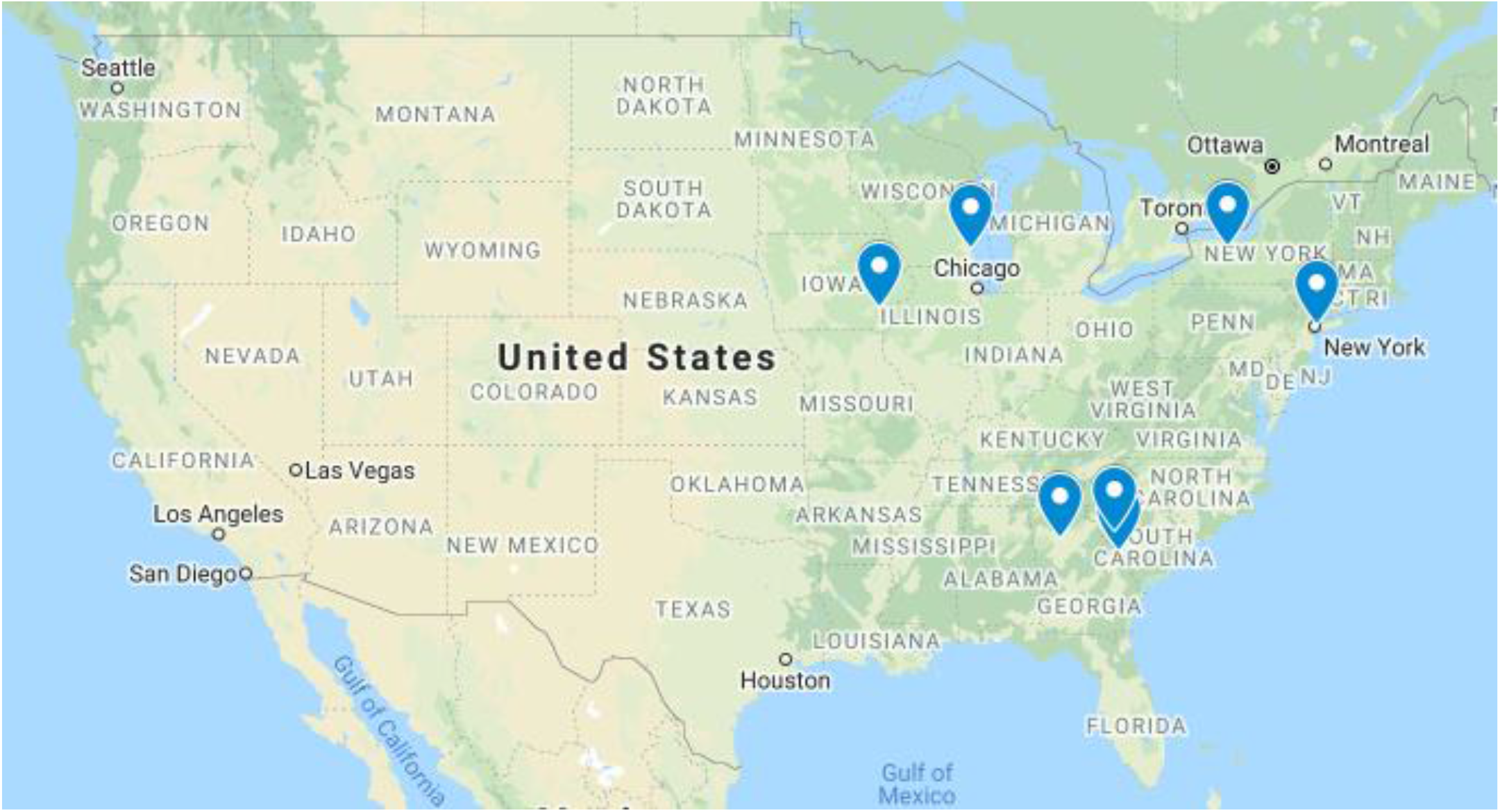
Site map of laboratory testing sites (Sites 1-5) in blue. Enrollment and consent of patients or waived authorization of de-identified, banked specimens was provided under the following IRBs: Biomedical IRB #00000150, Augusta University, Human Assurance Committee IRB # 611298, Wester-Copernicus Group IRB 20203726, The Self Regional Healthcare IRB numbers Pro00085001 and Pro00107951, Columbia University Human Subjects IRB AAAT9083/M00Y01. All PHI was removed and data were anonymized (coded and double-blinded) before accessing for the study.

## Methods

### Cohort design

This preliminary study includes 202 unique retrospective clinical research samples, aliquoted and frozen such that intra-run, inter-run, and inter-site runs could be assessed for a total of 331 sample runs. This is a double-blinded, de-identified, observational study approved through Biomedical IRB #00000150, Augusta University, Human Assurance Committee IRB # 611298, Western-Copernicus Group IRB 20203726, SRH IRB numbers Pro00085001 and Pro00107951, Columbia University Human Subjects IRB AAAT9083/M00Y01. All participants were consented or the need for consent was waived under the purview of the IRB guidelines. All samples were de-identified and all Protected Health Information (PHI) was removed before accessing for the study.

All samples in this retrospective study were previously tested with current SOC genetic testing methodologies and were grouped into three categories: 1) cases with a genetic diagnosis (n=152), 2) cases without a genetic diagnosis (n=6), and 3) controls (n=44). The diagnosed category included individuals that harbored a known pathogenic genomic variant (n=95 rare CNVs, n=38 FXS, n=8 FSHD1) or carrier genotype from prior clinical testing (n=11). The undiagnosed category included affected individuals *without* a known genetic diagnosis that could explain the clinical indications (negative or non-diagnostic results from CMA, karyotype, FISH, and/or other methodologies).

Certain SVs beyond the limitations of the OGM technology were excluded from this study, these are balanced Robertsonian translocations, balanced translocations with centromeric breakpoints, and other breakpoints in very large segmental duplications, and variants in mosaic cases below 20% fraction of cells with abnormality.

### Sample processing

Ultra-high molecular weight (UHMW) DNA was extracted, labeled, and imaged following the manufacturer’s protocol (Bionano Genomics Inc, USA) from peripheral blood collected in EDTA vials or cultured cells. Briefly, 650 μl aliquots of frozen blood samples or cultured cells in freezing media stored at -80°C were thawed immediately at 37°C in a swirling water bath for 2 minutes. The cells were counted using HemoCue WBC Analyzer or hemocytometer for blood and cultured cells, respectively. 1.5 million cells for blood samples and 1 million cultured cells were pelleted, homogenized in cell buffer, and digested with lysis buffer and Proteinase K. DNA was precipitated with isopropanol and bound with a nanobind magnetic disk. Bound UHMW DNA was washed using wash buffers A and B, resuspended in the elution buffer, and quantified with Qubit dsDNA assay kits (Thermo Fisher Scientific Inc. USA). DNA labeling was performed using Direct Labeling Enzyme 1 (DLE-1) with 750ng of purified UHMW DNA. Labeled DNA was loaded on Saphyr chips for imaging. The fluorescently labeled DNA molecules were imaged sequentially across nanochannel arrays on a Saphyr instrument. A target of 800Gbp was set for data collection for each sample.

### Assay quality control

UHMW DNA quality was assessed based on observation of viscosity homogeneity, and concentration measurements according to manufacturer guidelines. The first pass assay success rate, which includes DNA extraction, DLE1 labeling, and Saphyr chip run, was defined based on the output data meeting set QC thresholds with a single round of performing the assay. The final pass rate was defined based on output data meeting or being near the set QC thresholds, where one or more rounds of the assay were performed. The molecule quality report (MQR) was generated for each data set and included three key metrics that were used to evaluate sample QC: molecule N50, map rate (MR), and effective coverage. Molecule N50 was used to assess the size distribution of DNA greater than 150 kbp. The map rate metric was a calculation of the fraction of the DNA molecules that were able to align to the human reference (GRCh38). The effective coverage was calculated by the coverage depth of molecules that could be aligned to the reference genome (GRCh38). Analytical QC targets were set to achieve >160X effective coverage of the genome, 70% mapping rate, and 230 kbp N50 (of molecules >150 kbp).

Postanalytical quality control performance was determined by assessing the sex of the specimen and measurement of performance at stable regions of the genome. The EnFocus FXS analysis pipeline was used to infer samples’ sex, and pass/fail criteria for the following post-analytical quality metrics. The stable regions of the genome include one region from each autosome, which has been established to be stable in control populations. Based on expected sizing errors, the absolute percent differences between the OGM map and the reference should not exceed 1.2%. The pipeline requires that at least 90% of the stable regions be under this threshold [**15**].

### Data analysis

Genome analysis was performed using OGM specific pipelines, Bionano Access and Solve (versions 1.7 and 3.7, respectively), for data processing and variant calling. De novo assembly was performed using Bionano’s custom assembler software based on the Overlap-Layout-Consensus paradigm. A pairwise comparison of all DNA molecules was done to generate the initial consensus genome maps. Genome maps were further refined and extended with the best matching molecules. SV were called based on the alignment profiles between the *de novo* assembled genome maps and the Human Genome Reference Consortium GRCh38 assembly. If the assembled map did not align contiguously to the reference but instead was punctuated by internal alignment gaps or end alignment gaps, then a putative SV was identified. Fractional copy number (CN) analyses were performed from the alignment of molecules and labels against GRCh38. A sample’s raw label coverage was normalized against relative coverage from normal human controls, segmented, and baseline CN state estimated from calculating mode of coverage of all labels. If chromosome Y molecules were present, baseline coverage in sex chromosomes was halved. With a baseline estimated, CN states of segmented genomic intervals were assessed for significant increase/decrease from the baseline. Corresponding copy number gains and losses were exported. Aneuploidies are called using normalized read depth comparisons between chromosomes. At the same time, the absence of heterozygosity (AOH) is also called when larger than expected compared to the distribution in a control database, this tool will call AOH when it’s larger than 25 Mbp and in the whole sample (i.e. excluding mosaics).

Genomic variants consistent with FXS and facioscapulohumeral muscular dystrophy type 1 (FSHD1) were accessed using Bionano EnFocus pipelines [**15**]. For FXS analysis, the size of the CGG repeats was inferred based on the measured distance between two neighboring labels on the assembled map that contains the *FMR1* gene. By incorporating a known set of control map measurements, the pipeline estimates the most likely CGG repeat count with high/low repeat count (99% credible interval) and a probabilistic model estimating that the expansion exceeds the pathogenic threshold of >200 repeat units. For FSHD1 analysis, genomic DNA molecules aligning to regions of interest in chromosomes 4 and 10 were extracted and assembled. The resulting consensus maps were used to predict D4Z4 repeat size and assigned genotype of the permissive and non-permissive alleles (4qA and 4qB respectively). If enabled in Bionano Access, the pipeline added additional SVs and CN changes that were in proximity to the D4Z4 repeats on chromosome 4 (within 1 Mbp). CNVs overlapping the *SMCHD1* gene on chromosome 18 (which may impact chromatin remodeling and contribute to FSHD type 2) were also assessed.

Bionano Access was used for variant annotation and filtering. Variants were filtered based on defined guidelines with specific criteria including SV recommended confidence scores; size ≥1.5 kbp for insertions and deletions called by the de novo pipeline and ≥500 kbp for CNVs called by the read depth pipeline, frequency ≤1% population frequency (Bionano database of >300 healthy individuals); and SV Gene Overlap – set to include variants that overlap (within 3 kbp for SVs and within 100 kbp for CNVs) genes within GRCh38.

### SV Interpretation

SVs passing filtration steps outlined above were analyzed and classified into one of the following categories: pathogenic, likely pathogenic, variant of uncertain significance (VUS), likely benign or benign, according to ACMG guidelines [**22**]. SV analysis and classification was performed in Bionano Access, where each case was first analyzed by a laboratory analyst, and then the initial classifications were reviewed by a laboratory director. The laboratory directors also noted large-scale abnormalities (e.g. potential aneuploidies, regions of AOH, and triploidy) and some translocation events that were not yet integrated into the automated analysis result.

### Concordance Assessment

OGM data on processed samples were compared to their respective SOC test results, including sex and phenotype. SOC test results and clinical information was unblinded to a single individual for technical concordance assessment. Variant concordance was assessed by software calling, visual inspection, and manual curation. Concordance was only performed for the samples submitted and previously classified with SOC testing as pathogenic, likely pathogenic, and VUS. Concordance was also established for sample replicates.

## Results

### Cohort description

The data comprises of 331 datasets generated with the Saphyr system representing 202 unique patient samples (and replicates). Samples were run at five sites with CLIA certification (figure 1), including laboratory and medical directors with more than 175 years of combined experience with board certifications in the laboratory or medical genetics and/or pathology.

### Quality control performance

All samples passed pre-analytical requirements, including a cell count of at least 1.5 million cells per aliquot for blood samples and 1 million cells for cell cultures measured using a HemoCue WBC Analyzer and hemocytometer, respectively.

First pass performance was evaluated across five sites and and 93.8% of samples succeeded to meet data generation guidelines on the first round of lab work. Of the samples that needed to be repeated, all but three were successful, resulting in a final pass rate of 99.1% of all samples that could be analyzed for variants. Final analytical average QC metrics well exceeded the set targets: N50 (≥150 kbp) of 278.0 kbp, map rate of 88.4%, and average effective coverage of 223X. Of all samples, 98.1% had effective coverage ≥160x and 98.8% of samples met or exceeded 70% map rate. 92.1% passed the N50 target of ≥230 kbp and 99.7% had an N50 of ≥210 kbp (figure 2).

**Figure 2.**
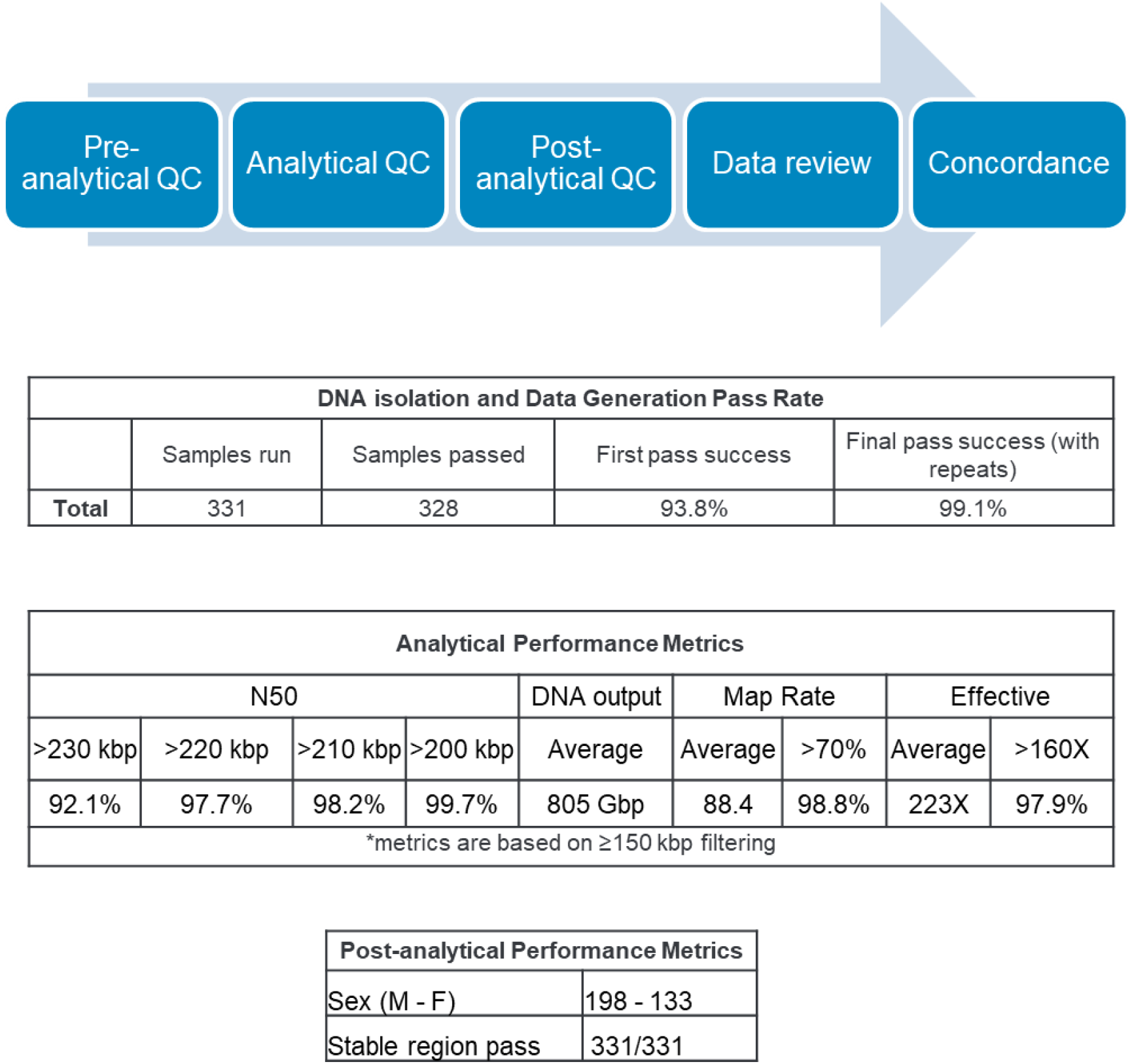
First pass success rate, analytical QC metrics, and post-analytical QC metrics. First pass success rate is defined as data meeting approximate QC standards with a single round of lab work (extraction, labeling, and chip flow cell). The final pass rate is defined as data meeting approximate QC standards in which one or more rounds of lab work were required. The results of analytical and post-analytical QC metrics as defined in the materials and methods section are shown.

All samples (100%) passed the post-analytical QC of stable region assessment. Post-analytical inference of sex determined 198 male and 133 female samples (figure 2).

### SV calls and SV filtering

SV calls in each sample were made by running the de novo assembly pipeline, approximately 800 Gbp of data was used from each sample to enable the detection of mosaic variants down to approximately 20% mosaic fraction. The average number of SVs detected is shown in table 1 with a total of 4266 variants per sample. These variants were filtered by a systematic filtering standard operating guideline (SOP) that was devised for this study. It includes selecting for large variants (insertions and deletions ≥1.5 kbp, CNVs from the coverage depth pipeline ≥500 kbp), identifying rare SVs in the human population (≤ 1% in the Bionano control sample SV database), finding variants overlapping or near genes (SVs within 3 kbp and CNVs from read depth analysis within 100 kbp). After filtering, there are only an average of 16.6 SVs per sample to be assessed for clinical significance.

**Table 1.** Average unique SV counts by type. The de novo assembly analysis pipeline generates annotated variants, which are then assessed in Bionano Access software v1.7. Here the SVs from a single sample are subjected to a baseline set of filters, which pass thousands of variants in an average sample and include polymorphic variants. Subsequently, an analyst can use the constitutional variant filtering guidelines to generate a relatively small number of variants that require review and classification, shown in the post-filter row. The workflow requires variants to be ≥1.5 kbp, nearly absent in a controls database (≤1% presence), and to overlap a gene within 3 kbp.

| SV Analysis Summary |  |  |  |
| --- | --- | --- | --- |
|  | Deletions | Duplications/<br>Insertions | Inversions/<br>Translocations |
| Baseline Average | 1259 | 2943 | 64 |
| Post Constitutional Filter<br>Average | 8.6 | 7.0 | 1.0 |

### Replicate analysis (including intra-run, inter-run, inter-site, and precision of variant positions)

As part of this study, 173 replicate samples from 47 unique cases were run to assess the reproducibility of the platform. Of these, 171 (98.8%) passed recommended map rate of >70% and effective coverage of 160x.

Among the cases with *FMR1* repeat expansions (> 200 CCG repeats), there were 63 replicates from 18 unique samples. Pathogenic expansions were detected in 63/63 specimens showing 100% concordance.

Additionally, 16 replicates from four unique cases with pathogenic SVs were also run. These samples had seven known pathogenic variants (three deletions, three duplications, and one translocation). OGM was 100% concordant in detecting all seven pathogenic variants in these replicates. The remaining replicates were from control samples with no known reportable variants and OGM results were 100% concordant.

### Technical concordance of OGM with SOC testing results

In total, 219 sample runs (including replicates) were evaluated for OGM concordance with the SOC tests. All classes of SVs were evaluated, including rare CNVs, balanced SVs, *FMR1* repeat expansions, and D4Z4 repeat contractions. Figures 3 and 4 show representative examples of each SV class. To date, 214/219 (97.7%) cases were found to be concordant and 5/219 (2.3%) were partially concordant (table 2 and S1). The five partially concordant samples each had multiple CNVs. In each case, one was classified as pathogenic and additional CNVs were classified as variant(s) of uncertain significance (VUS). Importantly, OGM detected the pathogenic CNV in each case but was unable to call the VUS findings in these cases. Possible explanations for these discrepancies are included in the discussion section below.

**Figure 3:**
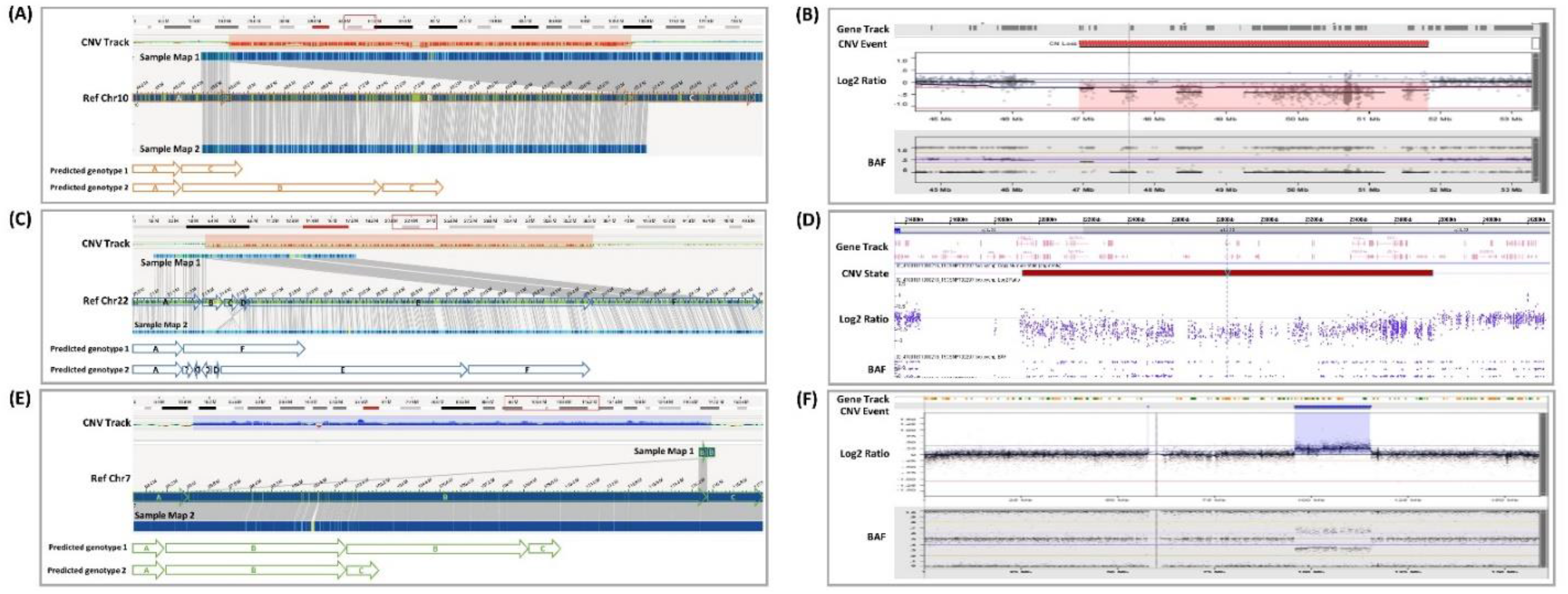
Representative examples of concordant microduplication and microdeletions. Panel A (top) shows a case with a 4.3 Mbp deletion at Chr 10q11.22q11.23 (chr10:45,794,475-50,135,024, hg38) identified and highlighted (red) in the CNV track. The deletion was also captured by Sample Map 1 generated by the de novo assembly algorithm. The bottom of panel A shows a diagram of the chromosomal structures predicted by the assembled maps illustrated by blocks with arrowhead indicating direction. Panel B shows the concordant CMA result of the Chr 10 deletion (chr10:46,900,000-51,800,000, hg19) from the same sample (visualized by NxClinical v.6.0 software, BioDiscovery, El Segundo, USA). A case with 22q11.2 LCR D-F (distal type I) deletion of 2.2 Mbp deletion (chr22:21312519-23542560, hg38) as shown in the CNV track and de novo assembly (Sample Map 1). In the bottom panel (C) LCR D-F deletion is illustrated by the loss of blocks B-E. A complex configuration of the LCR22 D segment was also noted in the predicted genotype 2. The CMA result (generated by ChAS software, ThermoFisher Scientific, Santa Clara, USA) (D) shows the concordant Chr 22q11.2 deletion (chr22: 21,922,619-23,654,064, hg19). A case with a 19.7 Mbp copy number gain of Chr 7q21.3q31.2 (chr7:96198684-115928803, hg38) is highlighted in blue, and the tandem duplication of this region is depicted by Sample Map 1 and illustrated in the bottom panel. The concordant CMA result of the Chr 7 duplication (chr7: 95,731,159-115,400,069, hg19; NxClinical v6.0 software, BioDiscovery).

**Figure 4:**
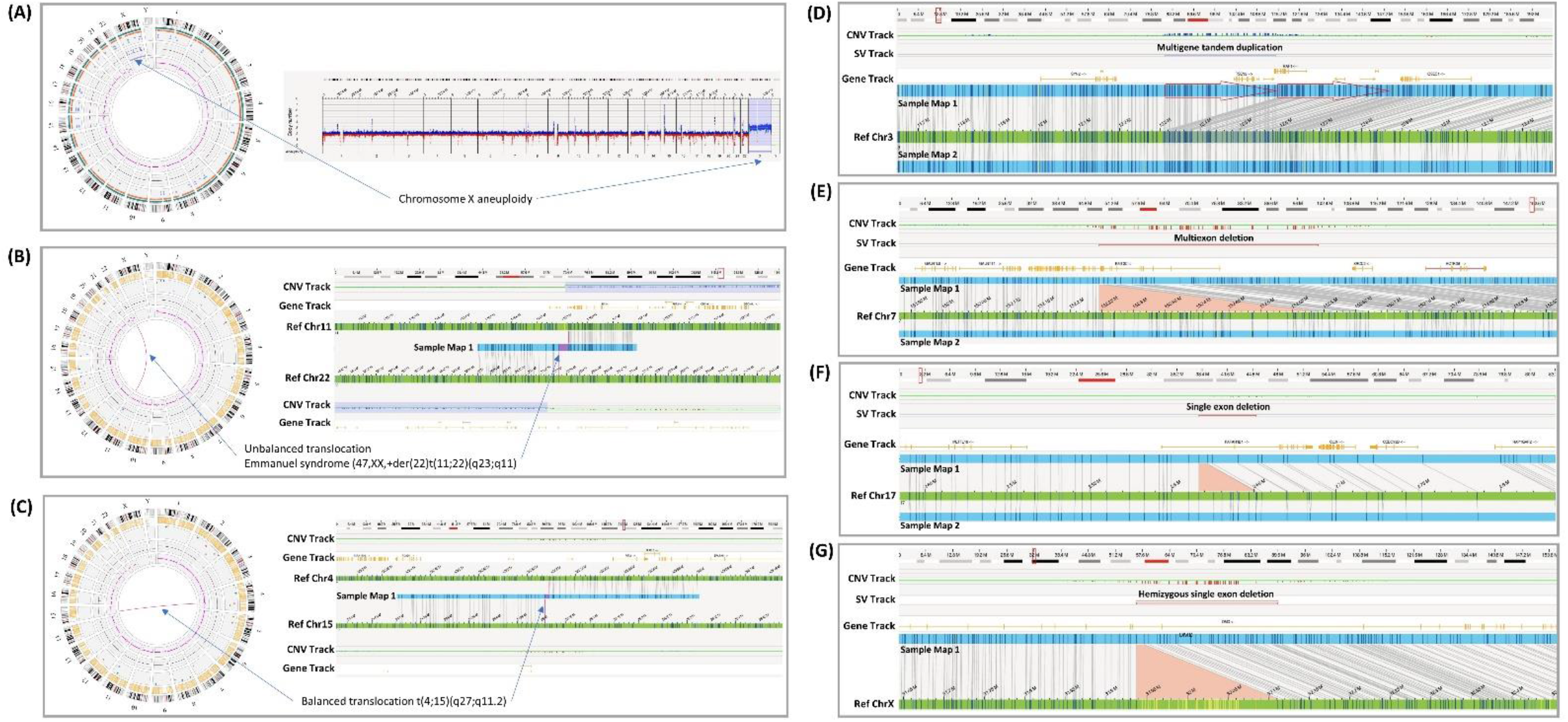
Representative examples of concordant SVs. A gain of Chr X is shown on the left panel, the right part of the panel shows the whole-genome view CNV track depicting the gain of Chr X (A). Rearrangement between chr11 and chr22 from an individual with Emmanuel syndrome (47, XX,+der(22)t(11;22)(q23;q11)) are shown in the circos plot, and in the right side of the panel, the unbalanced translocation was captured by Sample Map 1, also showing the copy number gain resulting from a der(22) chromosome (CNV tracks support the gains of chr11 and chr22) (B). A circos plot showing a balanced translocation detected between chromosomes 4q27 and 15q11.2 as indicated by a magenta line between chr4 and chr15, in the right side of the panel, the same translocation event was captured by Sample Map 1 (C). A 275 kbp tandem duplication (indicated by red arrows) was identified involving multiple genes including *PPARG, TSEN2, MKRN2*, and *RAF1* shown in Sample Map 1 (D). A 345 kbp intragenic deletion involving exons 1-14 of the *KMT2C* gene is shown in Sample Map 1 (E). A 29.8 kbp single exon deletion involving exon 2 of the *PAFAH1B1* gene was identified and indicated (red line) in the SV track (F). A 174 kbp hemizygous deletion involving exon 45 of the *DMD* gene is shown in Sample Map 1 (G).

**Table 2.**
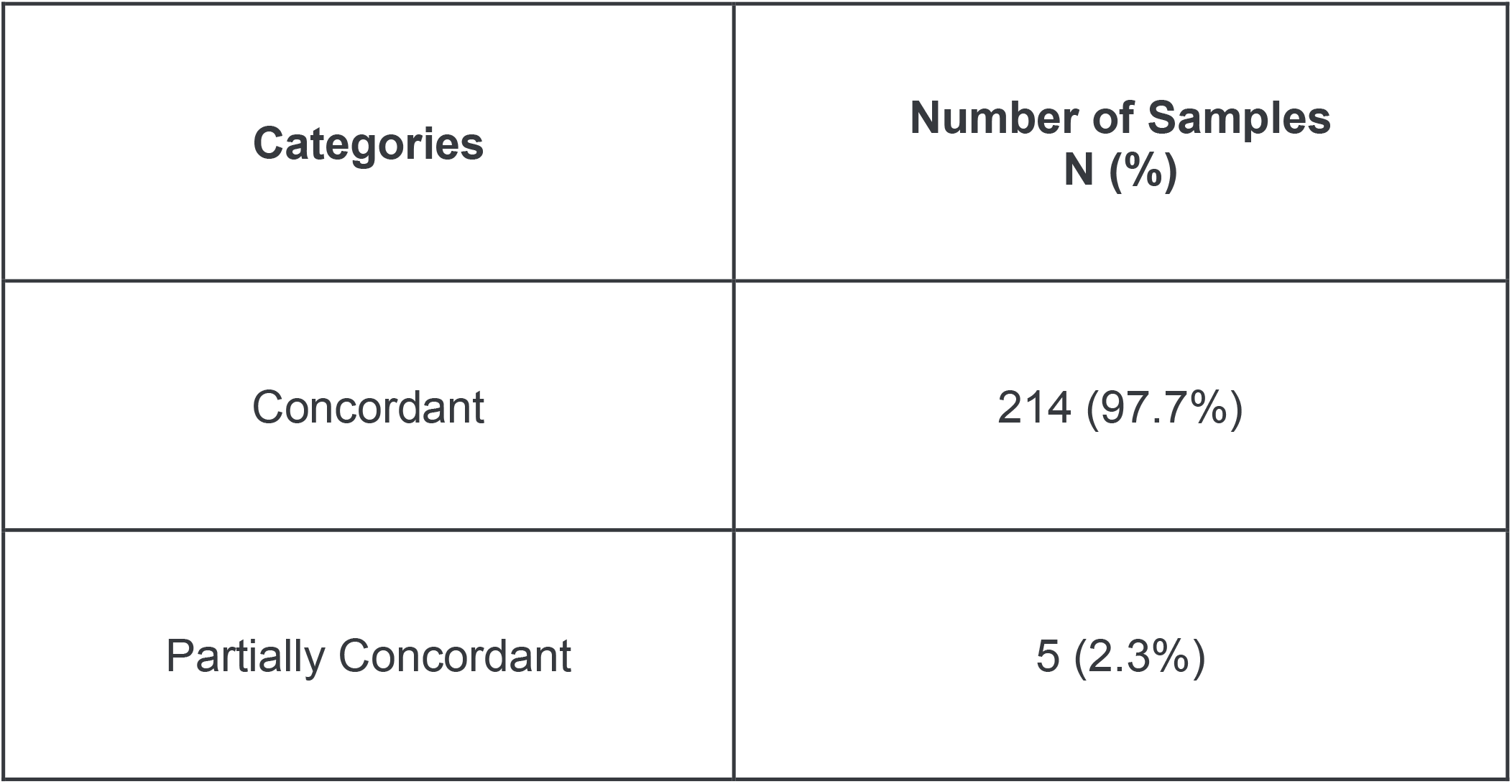
Technical concordance of OGM with SOC testing results. Pathogenic, likely pathogenic, and VUS variants detected by karyotype, FISH, and/or CMA were tested in this validation dataset. Samples were assessed for each variant by software calling, visual inspection, and manual curation. Details for this cohort can be seen in supplemental table S1.

| Categories | Number of Samples<br>N (%) |
| --- | --- |
| Concordant | 214 (97.7%) |
| Partially Concordant | 5 (2.3%) |

### Novel SV findings by OGM

In this preliminary study, one case which was previously negative by SOC testing had a novel insertion that could potentially explain the genetic cause of the clinical features in that individual. In a second case, OGM better defined the underlying structure of a likely pathogenic finding also identified by SOC testing. Both of these cases are described below.

*Case 1*. A child less than 5-years-old was referred for ES testing (which was non-diagnostic) due to neonatal seizures, epileptic encephalopathy/global developmental delay, failure to thrive, and acquired microcephaly. OGM detected a 269 kbp VUS duplication of Chr 13q14.3 (containing *SUGT1* and *CNMD* genes) and also detected a 5.9 kbp insertion in the *RORB* gene on Chr 9q21.13, potentially disrupting exons 5 and 6 (figure 5). *RORB* variants are associated with early-onset epilepsy, developmental delay/intellectual disability, and abnormal cortical response to photic stimulation (OMIM 601972) [**23**]. Parental studies are ongoing to assess the mode of inheritance of these SVs.

**Figure 5.**
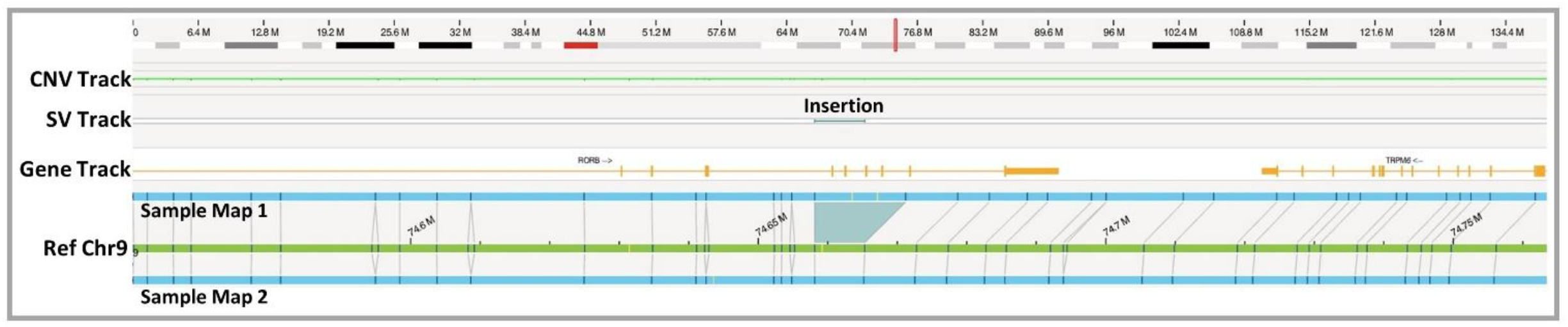
Representative Novel Finding by OGM: A 5.9 kbp insertion in the *RORB* gene is shown in a case with a neonatal-onset seizure disorder, cortical vision impairment, and developmental delay/intellectual disability. This insertion most likely disrupts the function of this gene. This sample was negative by SOC testing.

Case 2: A child between the ages of 6-10 years with ADHD, anxiety, and history of core weakness whose CMA revealed non-contiguous gains on Chr 1 {arr[hg19] 1p21.1p13.3 (104303487_109744359)x4,1p13.3(109749560_110114229)x3}. However, phasing (*cis vs. trans*) could not be elucidated from CMA data. OGM identified an intervening inversion indicating that the copy number gains represent a complex rearrangement [**24**].

## Discussion

ACMG, AAP, and AAN recommend genetic testing for the diagnosis of constitutional disorders, which include birth defects, pediatric, and adult neurodevelopmental disorders. A precise genetic diagnosis facilitates comprehensive clinical management of the patient and recurrence risk counseling for the family. In most cases where genotype/phenotype associations are non-informative, a tiered-genetic testing approach is required to end the diagnostic odyssey of individuals affected with rare genetic disorders [**25**]. Since the past decade, CMA has been employed as the first-tier test for genome-wide CNV analysis of individuals with developmental delay, intellectual disability, autism spectrum disorder (ASD), dysmorphic features, and multiple congenital anomalies [**5**]. Also, FXS testing is performed in conjunction with CMA testing in males with intellectual disability and/or autism [**11**]. In cases that have multiple CNV findings by CMA, karyotyping is recommended to ascertain structural rearrangements involving chromosomal translocations. Even though CMA has been demonstrated to have a higher diagnostic yield (∼20%), CMA has significant limitations when it comes to detecting balanced SVs (translocations and inversions), assessing the orientation of duplications, and low-level mosaic events (<20%). The methods used for sizing large repeat array expansions are laborious, time-consuming, and have low accuracy (southern blotting) or low dynamic range (PCR, and NGS). Also, a diagnostic yield of *FMR1* CGG repeat expansion analysis is reported to be 1-2% among patients with an intellectual disability or global developmental delay [**10**]. Interestingly, a diagnostic yield of ∼10% for CMA and ∼0.5% for *FMR1* CGG repeat analysis has been reported in patients with primarily autism spectrum disorders (ASD) [**10**]. In cases with a non-informative or negative result by CMA, ES is currently performed as the tier 2 test and has shown a diagnostic yield of ∼35% and ∼15% in patients with an intellectual disability or global developmental delay, and ASD, respectively [**12**]. It is evident that the current standard-of-care genetic testing workup includes several cytogenetic and molecular technologies to reach a conclusive genetic diagnosis, which is often a tedious and time-consuming process, causing significant economic and emotional burden to the patient and their families. Despite these challenges, a significant percentage of cases remain genetically undiagnosed.

OGM has shown the potential to consolidate multiple SOC methods due to its ability to detect several classes of SVs with high sensitivity and specificity (table 3). Recently, published studies have demonstrated 100% concordance of OGM with SOC methods [**20, 21**]. However, the technical performance of the assay across multiple sites with large datasets have not been assessed to date. In this study, 331 samples representative of diverse constitutional disorders with varying clinical indications and chromosomal abnormalities have been tested by OGM. The analytical validity and technical performance of OGM was assessed across 5 different testing laboratories in the US. The robustness of OGM is demonstrated by 99.1% of all samples successfully delivering a usable result across all sites. The first pass success rate, requiring no repreparations, was 93.8%, across all sites. For any new/innovative technology to be adopted for clinical or research use, it is imperative that appropriate performance indicators are used to monitor and assess the performance of the test/method. The following key analytical QC metrics for OGM were assessed: target map rate of molecules and effective (mapped) genome coverage was set to 70% and 160X, respectively, and these targets were achieved with 98.8% and 97.9% of samples, respectively (figure 2). In addition, molecule N50, an important measure of OGM’s ability to map and detect complex, genome-wide SVs was >220 kbp for 97.7% of samples (figure 2). In a CLIA setting, post-analytical QC metrics allow for assessing the overall performance and success of the assay, here we have used a pass/fail criteria to assess 22 stable regions of the genome (one region per autosome). 100% of the tested samples (331/331) passed the post-analytical QC threshold.

**Table 3.**
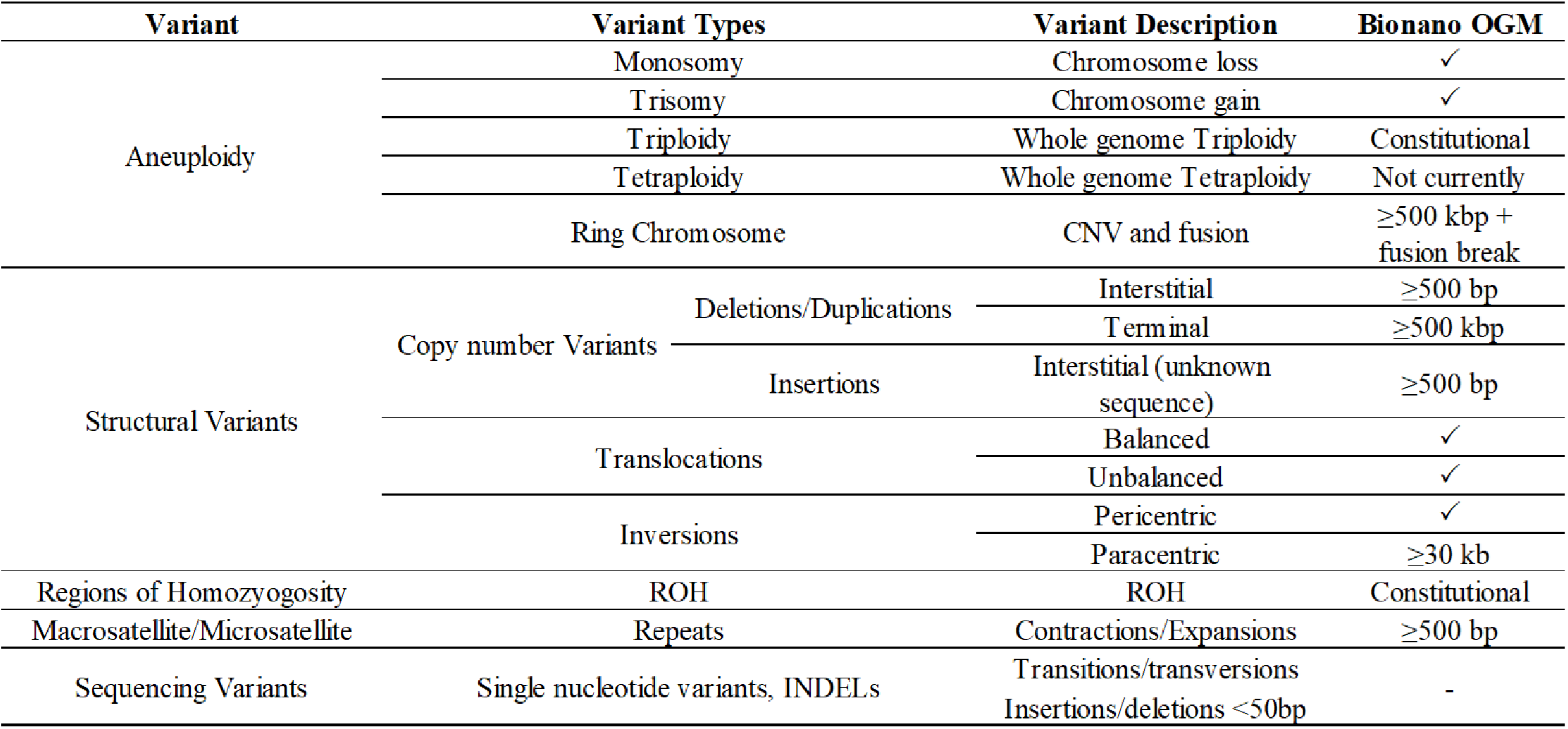
Variant classes. List of variant classes and OGM performance across each class.

The technical concordance of OGM in this multi-site, multi-operator and multi-instrument study was 97.7% (214/219) in detecting SVs compared to SOC methods. An important metric that was assessed in this study was OGM’s ability to detect all classes of SVs with a resolution of 500 bp. Our dataset reveals an average of 4266 unique SVs per sample, which is in agreement with published studies [**20**].

The aim of this study demonstrated the sensitivity of OGM in detecting several classes of SVs, CNVs, and repeat array analysis, and also including complex rearrangements that were previously detected using multiple SOC methods. The use of multiple bioinformatic tools in parallel for the detection of all different classes of SVs and AOH with a single dataset reinforces the benefit of implementing OGM in a clinical setting. The de novo analysis pipeline that assembles the genome into consensus maps and aligns with the reference assembly showed the reproducibility of the SVs detected. Although the resolution of this pipeline enables the detection down to 500 bp, the smallest known SV in our cohort was 23 kbp, previously reported using CMA. Additionally, the same pipeline also enabled the detection of repeat expansion and contractions in the genome thereby eliminating the need for multiple tests performed in a tiered approach to reach an accurate diagnosis. EnFocus analysis is a set of focused analyses enabling faster analysis and more specific and robust performance and faster analytical metrics for cases that had a clinical suspicion or indication of FXS and FSHD1. Another benefit of this assay was a second coverage-based algorithm that computes the copy number state and is analogous to CNV assessment tools used for CMA analysis. This pipeline allowed for the detection of large CNVs, aneuploidies and ROH across the genome with high confidence and ease.

Although FXS testing is recommended for individuals with developmental delay/intellectual disability [**11**], diagnostic yield is only 0.5-2% [**10**]. Considering that CMA is also recommended and often done in parallel with FXS testing, it would be highly advantageous to combine FXS testing with SV testing even if borderline expansions cannot be precisely defined and require infrequent reflex testing. OGM’s EnFocus tool was able to provide *FMR1* measurements and % likelihood of an expansion >200 CCG triplet repeats, and this allowed the users to rule in or rule out a pathogenic expansion. A separate study is being performed to assess the FXS test results when the expansions are near the threshold length (i.e. 150-250 repeats), in which case, a reflex PCR test might be needed.

The investigators of this multi-site study developed an efficient variant filtration protocol that reduced the number of variants to be analyzed to a manageable size of 16.6 per sample, thus, filtering out 99% of the polymorphic variants. The 98% technical concordance achieved in this double-blinded analysis compared to SOC methods shows the sensitivity and accuracy of the OGM assay and the data analysis protocol thereby instilling confidence that OGM can be implemented for clinical testing. The current software version of Bionano Access v1.7 enables the categorization of variants into ACMG classification criteria (pathogenic, likely pathogenic, VUS, likely benign, and benign) allowing analysts and lab directors to manage their internal database of SVs. Also, the analysis of each case was facilitated by the ability to use several readily available tools such as disease loci/gene bed files overlaying SV calls, Bionano internal population frequency database, preferred gene variant, DGV and UCSC links, in an integrated visualization software to efficiently assist the analysis team for variant classification.

Our technical concordance dataset revealed five samples (supplemental table S1), so far, that were considered partially concordant. The first is a putative duplication reported by CMA on Chr 2q14.2 (BNGOKH-0000141) and Chr 3q12.2 (BNGOKH-0003215); however, these copy gains were not readily visible within the assembled consensus genome maps of OGM. There are two plausible reasons for this discrepancy: 1) it is possible that the duplicated material is inserted into a different location of the genome and doesn’t possess a sufficient number of labels to accurately map its location 2) the duplication is present in only a subset of molecules in insufficient quantity to be assembled into a separate contig. It is important to note that while CMA detected the dosage of interrogated regions of the genome, it cannot assess the genomic locations or orientation of the duplications. Additionally, three samples (BNGOKH-0003205, BNGOKH-0003215, BNGOKH-0003245) possessed similar copy number losses on Chr 15q11.2 identified by CMA; however, these deletions are located close to the centromere where consensus genome maps generated by OGM are partially masked due to incomplete reference sequence as well as the possibility of genome map miss-assembly and alignment issues in repetitive regions. Similarly, for case 55752101092154_KH54-2, the identified copy number loss by CMA is located at the telomeric region of Chr 19p13.3 and also has high molecule alignment noise parameters that are typically masked.

This publication is the first of a series expected from expansion of this large dataset on OGM in 2022. The assay runs of additional samples (across all sites) are ongoing and are being extended to additional undiagnosed cases as well as samples with difficult to interpret results. We anticipate the full study (>500 samples) will robustly characterize the ability of OGM for the detection of variants missed by SOC testing and add informative SV findings in a subset of undiagnosed cases, as reported by Shieh *et al*. [**26**]. In one undiagnosed case OGM found a potentially relevant SV (associated with the phenotype of the affected individual) and in a second case OGM refined the breakpoints and architecture of the complex rearrangement.

The performance for samples with varying genomic aberrations reported for this study demonstrates that OGM offers the opportunity to avoid multiple testing modalities that add time and cost to any diagnostic workup. Two pediatric cases support the assertion that OGM could be a better diagnostic tool: the first case is a child who underwent several rounds of SOC testing including ES with no findings. In this case, OGM identified a putative diagnostic finding: an insertion potentially disrupting the *RORB* gene. Loss of function of this gene has been reported to cause early-onset seizures, developmental delay/intellectual disability, and cortical visual abnormalities – all of which are consistent with the clinical presentation of this patient [**23**]. If this case had an OGM test the families’ diagnostic odyssey may have ended several years earlier. A similar ability to improve diagnostic yield in postnatal cases has been shown in previous studies using OGM [**27**]. In a second case, a pediatric patient with a neurodevelopmental phenotype had two non-contiguous CNVs on chromosome 1 that were deemed likely pathogenic by CMA testing. In the same assay, OGM detected both CNVs as well as an intervening inversion that adds both clinical and reproductive detail that may be important for this child’s care and the family’s recurrence risk counseling in the future [**24**]. These examples from our cohort demonstrate the great potential of OGM as a more precise genomic diagnostic tool. Overall, OGM provides a simplified and streamlined workflow that can efficiently end the diagnostic odyssey of individuals with rare diseases.

## Conclusions

SVs, including CNVs, have been investigated for the genetic diagnosis of individuals affected with constitutional disorders by traditional cytogenetic methods, including CMA. On the other hand, sequencing-based assays are refractory to the detection of many structural rearrangements due to the repetitive nature of the human genome. The results from this study demonstrate the high technical performance of the OGM workflow from DNA isolation through data analysis. The inter and intra run replicate performance also demonstrate the reproducibility of the OGM technique so that it can be easily adapted and validated as a laboratory-developed test by CLIA laboratories. The novelty of OGM is not limited to CNV analysis alone (∼100% concordance with CNVs detected by CMA), but also in its ability to resolve balanced structural rearrangements, size *FMR1* repeat expansions and D4Z4 repeat contractions. Thus, a single assay like OGM allows the genetic laboratories to provide a rapid result to diagnosis besides reducing the economic burden on the affected individuals and their families. Further expansion of the dataset presented in this study is ongoing and collaboration between researchers, clinicians and families is underway to demonstrate the maximum benefit of the OGM technique.

## Data Availability

All relevant data has been made available in the manuscript and supplementary file.

## AUTHOR CONTRIBUTIONS

Conceptualization: BL, NS, PN, AB, AI, GS, UB, RS, SS, RK Methodology: BL, NS, PN, AB, AI, GS, UB, RK

Data curation: BL, NS, PN, AB, AS, AI, KA, GS, UB, RK

Writing, reviewing, editing: BL, NS, PN, AB, AI, GS, UB, RK Visualization: BL, NS, PN, AB, AS, AI, KA, GS, UB, RK

All authors have read and agreed to the published version of the manuscript.

## ACKNOWLEDGEMENTS

We wish to thank the Greenwood Genetic Center, their clinical testing laboratory, and their affiliated physicians for their participation and clinical acumen; without their expertise, many of these diagnoses would not have been made to enable the validation of new genomic technologies. In addition, we thank several other groups who assisted in patient recruitment for known genomic findings, including the Facioscapulohumeral Muscular Dystrophy Support Group, the 4p-Society, the 5p-Society, the Balanced Translocation Support Group, and the University of Utah Maternal-Fetal Medicine Center. We also acknowledge Baylor’s Undiagnosed Diseases Network—and especially Jill Rosenfeld Mokry, CGC—for assisting in patient recruitment and ongoing discussion. We would like to thank the technical staff at each of the participating sites. We express gratitude to the children and families who submitted samples (which are often difficult to collect) to further genomic discovery.

## ETHICS DECLARATION

The study was conducted according to the guidelines of the Declaration of Helsinki and approved by the following Institutional Review Boards: A-BIOMEDICAL I (IRB registration #00000150), Augusta University; Human Assurance Committee IRB # 611298; Western IRB – Copernicus Group (WCG), study number 20203726, (University of Rochester Medical Center, Medical College of Wisconsin, Columbia University Medical Center, Greenwood Genetic Center, Augusta University, Praxis Genomics, University of Iowa Health Clinics); the Self Regional Healthcare IRB numbers Pro00085001 and Pro00107951 (University of Rochester Medical Center, Medical College of Wisconsin, Columbia University Medical Center, Greenwood Genetic Center, Augusta University, Praxis Genomics, University of Iowa Health Clinics); and the Columbia University Human Subjects IRB, protocol number AAAT9083/M00Y01. This includes informed consent provided by individuals with newly collected samples or waived authorization for use of de-identified, banked samples. All protected health information (PHI) was removed and data were anonymized (coded and double-blinded) before accessioning for the study.

## Supplementary material

**Supplemental Table S1. ISCN nomenclature** Table of variants assessed for concordance with SOC testing in ISCN format.

